# Collective Impacts of Demographic and Clinical Factors on Psycho-emotional States of Patients with Hepatocellular Carcinoma: A Cross-Sectional Study

**DOI:** 10.1101/2023.12.30.23300609

**Authors:** Crystal Chia-Chun Liang, I-Ju Pan

## Abstract

**Background:** To appraise comprehensively the demographic and clinical factors that may collectively contribute to demoralization, depression and negative meaning of life in patients with cancer, employing hepatocellular carcinoma (HCC) as the representative investigative subject.

**Purpose:** To appraise comprehensively the demographic and clinical factors that collectively contribute to demoralization, depression and negative meaning of life in patients with cancer, employing hepatocellular carcinoma (HCC) as the representative malignancy for investigation in a Taiwanese population.

**Design:** A retrospective cross-sectional study from 2019–2020 on 250 HCC patients in hospitalized and following the STROBE guidelines.

**Methods:** Prevalence was estimated by G-power; and associations between demographic or clinical factors and scores of Mandarin versions of Demoralization Scale (DS-MV), Center for Epidemiologic Studies Depression Scale (CES-D) or Meaning in Life Scale (MiLs) were evaluated by Chi-square test or Pearson’s product-moment correlation.

**Results:** Factors with DS-MV scores higher than 30 included marital status, children as primary caregiver, gender and metastasis. Factors with CES-D scores higher than 16 included marital status, children as primary caregiver, age, monthly financial support and cancer comorbidity. Factors with MiLs scores less than 7 included marital status, and recurrence or duration of cancer.

**Conclusions:** Marital status is a major contributing factor to all three psychological states. Children as primary caregiver impacts more critically than spouse on both demoralization and depression; gender and metastasis are akin to demoralization; age, monthly financial support and cancer comorbidity contribute to depression; and recurrence and duration of cancer impact on negative meaning of life.

**Impact:** This is first time that collective demographic and clinical factors on the mental health of patients with liver cancer are identified. Our findings would encourage clinical nurses to be more conscious of the contribution of these factors on the emotional states of patients with cancer when performing their daily duties.

## 1. INTRODUCTION

With the advances in contemporary medicine that allow for early diagnosis and better treatment strategies, the five-year survival rate of patients with malignant tumors has increased from 17.8% to 21.4% (Roth et al., 2018). Because of the improved prognosis and the resultant chronic nature of malignancy, the mentality of some patients with cancer may shift from waiting for death to pursuit of quality life. Other patients who are unable to coexist peacefully with cancer, however, may face psychological problems, including demoralization and depression, and ponder the meaning of life. The continuous feeling of powerless and helpless, along with failure to receive timely support, may lead an individual to hopelessness and isolation, which will eventually affect personal values, meaning of life and purpose (Vehling et al., 2012).

## 2. BACKGROUND

Demoralization is a feeling of helplessness, hopelessness, meaningless, subjective incapacity and loss of self-esteem when an individual is unable to cope with a specific event and becomes ill-adjusted (Clarke & Kissane, 2002; de Figueiredo, 1983). Living alone, poor physical conditions and location of tumors are factors reportedly related to manifestation of symptoms of demoralization in young females, although marriage, religion and cancer stage appear to be unrelated (Fang et al., 2014; Ko et al., 2018; Li et al.,2017). Patients with good family support and high degree of education are less likely to experience demoralization. Regardless of the type of malignancy or geographic location, the rate of demoralization among patients with cancer ranges from 13% to 49% (Ko et al., 2018; Tang et al., 2020, Wang et al., 2023).

Depressive symptoms may develop when an individual who faces a stressful event is unable to cope or copes poorly. These symptoms include negative perception of oneself, negative self-esteem, erroneous logical thinking, exaggeration of negative events in life, reduced interaction with the outside world, and hopelessness and pessimism about the future (Beck, 1997; Kendall et al., 1987). The proportion of depressive symptoms among patients with cancer is about 12.9% to 35%. Female patients, and those younger than 50 years, less educated or with other health complications exhibit higher levels of depression. (Gonzalez et al., 2014; Kim et al., 2016; Tabriz et al., 2019).

From satisfying basic needs to pursuing meaningful and realistic existence, humans strive to achieve consistent goals in their lives. These goals are further expanded by the combination of individual experience and societal environment to create enriched and diverse values. The pursuit and realization of valuable goals and the accompanied satisfaction therefore dictate the meaning of life (Park & Folkman, 1997). Whereas strong societal support, higher education level, and longer duration of cancer are positively related to the meaning of life, anxiety and poor financial support exert negative impact (Elekes, 2017; Jim & Andersen, 2007; Vehling et al., 2012).

Individual factors that contribute to demoralization, depression and meaning of life in patients with cancers have been reported (Elekes, 2017; Ko et al.,2018; Li, Ho, Wang.,2017; Su, 2018; Vehling et al., 2012). For example, age, gender, and physical condition of patients with cancer are contributory factors to demoralization and depression, but gender and age have no significant influence on the meaning of life. Provision of care to patients with cancer also depends on ethnic and cultural backgrounds. Nevertheless, a comprehensive investigation on the collective contribution of demographic (age, gender, marital status, education, monthly financial support, occupation, religion and caregiver) and clinical factors (cancer stage, cancer recurrence, cancer metastasis, cancer duration, cancer comorbidity and treatment schemes) in accordance with Asian practice of caregiving on demoralization, depression and meaning of life in patients with cancers is still lacking.

According to World Health Organization (WHO) statistics, the number of deaths due to liver cancer (hepatocellular carcinoma, HCC) in the world in 2019 is 577,000, which ranks fifth in global cancer deaths (WHO-Global health estimates: Leading causes of death, 2021). In Taiwan, malignant tumor places first on the top ten causes of death in 2021. With 34.0 patients per 100,000 population, liver cancer ranks second among all cancers (Ministry of Health and Welfare, 2023). Employing HCC as the representative investigative subject because of its prevalence in Taiwan, together with a quantitative research design, the present study evaluated the collective demographic and clinical factors that may contribute to demoralization, depression and meaning of life in patients with cancer. We also explored the influence of the main caregiver on the psychological state of patients based on care practice according to Asian culture, as exemplified by the custom in Taiwan.

## 3. METHODS

### 3.1 Study Design, Setting and Ethical Approval

This study employed a retrospective cross-sectional design, and was carried out in 2019–2020 at a medical center located in southern Taiwan. Ethical approval was obtained from the Institutional Review Board (IRB number: 201901213B0) and from the data protection officer at the hospital where the patients were recruited. All participants provided informed written consent and were guaranteed confidentiality and the right to withdraw from the study at any time. The patients also gave their permission to use their data obtained from the medical records of the hospital. The study adhered to the Strengthening the Reporting of Observational Studies in Epidemiology (STROBE) guidelines for cohort, case-control and cross-sectional studies.

### 3.2 Study population

The study sample was drawn from the electronic hospital records of individuals treated during 2019–2020 for HCC with curative intent. The inclusion criteria were: patients diagnosed with liver cancer; between 20–80 years of age; agreed to participate in the research program; willing to sign the informed consent; able to communicate in spoken or written Mandarin and free from mental illness. The exclusion criteria were: patients not diagnosed with HCC; under 20 or over 80 years of age; unable to communicate effectively; diagnosis of mental or intellectual disability and failure to sign the informed consent.

### 3.3 Data collection

The data were collected from patients hospitalized between September 2019 and September 2020. A questionnaire for demographic information (age, gender, marital status, education, monthly financial support, occupation, religion and caregiver) was developed for this study. Clinical data (cancer stage, cancer recurrence, cancer metastasis, cancer duration, cancer comorbidity and treatment schemes) were collected from the electronic hospital records. The purpose and study design were explained to the participants before they completed the questionnaire and signed the informed consent.

### 3.4 Measures

#### 3.4.1 Demoralization Scale

The Demoralization Scale (DS) was developed by Kissane DW in 2004 based on the definition of demoralization by Engel and Frank. DS can be used to assess the severity of demoralization, predict suicidal intention, evaluate quality of life and screen patients with cancer who do not meet the diagnosis of depression but face survival crisis and lose meaning of life. The mandarin version of DC (DS-MV) was used in this study. With the Cronbach’s alpha value of the total scale at .92, the reliability and validity of DC-MV were confirmed by Li et al. (2018b). Based on a Liker scale of 0 to 4 point for each of the 24 items, a score of 30 or above over a range of 0-96 points was considered high level of demoralization (Fang et al., 2014). The Cronbach’s alpha value of the scale in this study was .91.

#### 3.4.2 Center for Epidemiologic Studies Depression Scale

This study employed the Mandarin Version of the Center for Epidemiologic Studies Depression Scale (CES-D) published by Randloff in 1977, as translated by Chien and Cheng (1985), to evaluate the depressive symptoms of the subjects. With the Cronbach’s alpha value of the total scale at .85, the reliability and validity of CESD were confirmed by Randloff (1977). Based on a Liker scale of 0 to 3 points for each of the 20 items, the total score ranges from 0 to 60 points. A score greater than 16 was indicative of a tendency to be depressed; the higher the score, the more prone to depression (Radloff, 1977). The Cronbach’s alpha value of the scale in this study was .88.

#### 3.4.3 Meaning in Life Scale

This study used the mandarin version of Meaning in Life Scale (MiLs) originally developed by Jim et al. (2006) that we translated, which is made up of 21 questions on the impact of cancer diagnosis and treatment on life. Over a range of −3 to 17 as a result of subtracting the scores of the positive questions from the negative questions, a score of 7 points was used to distinguish positive from negative meaning of life. The Cronbach’s alpha value of the original scale is .93 (Jim & Andersen, 2007); the Cronbach’s alpha value of the scale in this study was .8.

### 3.4 Sample size calculations

Based on the assumption that the statistical result is a medium effect size (Cohen, 1988), 250 cases are required to achieve a significance level of .05 and a statistical power of .80. At a sample loss rate of 15%, the number of participants to be recruited for this study should be at least 285.

### 3.5 Data analyses

All data were analyzed using SPSS version 25 (IBM Corp. Released, 2019) descriptive statistical analysis. Demographic and clinical data were shown in number and percentage; DS-MV, CES-D and MiLs scores were compared by Chi-square test if expected values were less than 5 used Fisher’s exact test. Pearson’s product-moment correlation was used to analyze the correlation between the continuous variables (age and cancer duration) and DS-MV, CES-D or MiLs in patients with liver cancer, and the correlation of the DS-MV, CES-D and MiLs scores.

## 4. RESULTS

### 4.1 Demographic and clinical variables

Of 290 eligible patients with liver cancer, 250 agreed to sign the written consent and the survey inventory, and completed the questionnaires, resulting in a response rate of 84%. 28 patients refused to participate after the purpose and study design were explained to them, and 12 participants were unable to sign the consent because they could not read or write.

As presented in **Table 1**, 48.4% aged between 61 to 70 years, 83.6% participants were male, 78.8% were married, 50.4% received middle and high school education, 93.6% had no economic problems, 35.6% were retired, and 60.8% had their spouse as the main caregivers during hospitalization. Cancer recurrence was 51.2%, 94.0% had no cancer metastasis and 70.0% patients exhibited no physical comorbidity from liver cancer.

**TABLE 1.** Participant characteristics and scales of study and sample (N=250)

| Variable | n (%) | Mean ± SD | Variable | n (%) | Mean ± SD |
| --- | --- | --- | --- | --- | --- |
| Age | | 60.9 ± 8.7 | Cancer stage | | |
| ≤ 50 | 32(12.8) |  | BCLC 0 | 32(12.8) |  |
| 51-60 | 69(27.6) |  | BCLC A | 90(36.0) |  |
| 61-70 | 121(48.4) |  | BCLC B | 98(39.2) |  |
| >71 | 28(11.2) |  | BCLC C | 30(12.0) |  |
| Gender |  |  | Cancer recurrence |  |  |
| Male | 209(83.6) |  | No recurrence | 122(48.8) |  |
| Female | 41(16.4) |  | Recurrence | 128(51.2) |  |
| Marital Status |  |  | Cancer Metastasis |  |  |
| Single | 53(21.2) |  | No | 235(94.0) |  |
| Married | 197(78.8) |  | Yes | 15(6.0) |  |
| Educational level |  |  | Cancer Comorbidity |  |  |
| Elementary and below | 70(28.0) |  | No | 175(70.0) |  |
| Middle and High school | 126(50.4) |  | Yes | 75(30.0) |  |
| College above | 54(21.6) |  | Treatment schemes |  |  |
| Monthly financial support |  |  | Surgical | 57(22.8) |  |
| Enough | 234(93.6) |  | Medical | 109(43.6) |  |
| Not enough | 16(6.4) |  | Other | 84(33.6) |  |
| Occupation | | | DS-MV | | 27.8 ± 8.56 |
| None | 89(35.6) |  | <29 | 184(73.6) |  |
| Industry | 66(26.4) |  | ≥30 | 66(26.4) |  |
| Business | 42(16.8) | | CESD | | 7.69 ± 7.40 |
| Others | 53(21.2) |  | ≤15 | 219(87.6) |  |
| Caregiver |  |  | >16 | 31(12.4) |  |
| None or Caretaker | 44(17.6) | | MiLs | | 8.63 ± 2.00 |
| Spouse | 152(60.8) |  | <7 | 53(21.2) |  |
| Children | 54(21.6) |  | ≥7 | 197(78.8) |  |
| Religion |  |  |  |  |  |
| None or others | 41(16.4) |  |  |  |  |
| Buddhism | 55(22.0) |  |  |  |  |
| Taoism | 154(61.6) |  |  |  |  |

Abbreviations: DS-MV= Mandarin versions of Demoralization Scale; CES-D= Center for Epidemiologic Studies Depression Scale; MiLs= Meaning in Life Scale

### 4.2 Correlation between demoralization, depression and negative meaning of life in HCC patients

To set the stage of our study, Pearson’s correlation was used to analyze the relationship between demoralization, depression and negative meaning of life in our patients. The results showed that total score of DS-MV was positively correlated with total score of CES-D (*r*=.53, *p*<.01). Total score of DS-MV and total score of MiLs were negatively correlated (*r*=-.52, *p*<.01), as were total score of CES-D and total score of MiLs (*r*=-.35, *p*<.01) **(Table 2)**.

**TABLE 2.** Correlation between demoralization, depression and negative meaning of life in HCC patients (N=250)

| Variable |  | DS-MV | CES-D | MiLs |
| --- | --- | --- | --- | --- |
| DS-MV | Pearson | 1 |  |  |
| CES-D | Pearson | .53** | 1 |  |
| MiLs | Pearson | -.52** | -.35** | 1 |

Abbreviations: DS-MV= Mandarin versions of Demoralization Scale; CES-D= Center for Epidemiologic Studies Depression Scale; MiLs= Meaning in Life Scale
\* $p < .05$ , \*\* $p < .001$

### 4.3 Relationship between demographic factors and demoralization, depression or negative meaning of life in HCC patients

Chi-square test was used to compare the dichotomous differences between demographic data and demoralization, depression or negative meaning of life in patients with HCC **(Table 3)**. There was no significant difference between education level, occupation or religion and DS-MV, CES-D or MiLs, nor between monthly financial support and DS-MV (*χ^2^*=1.08, *p*=.38) or MiLs (*χ^2^*=1.03, *p*=.34).

**TABLE 3.**
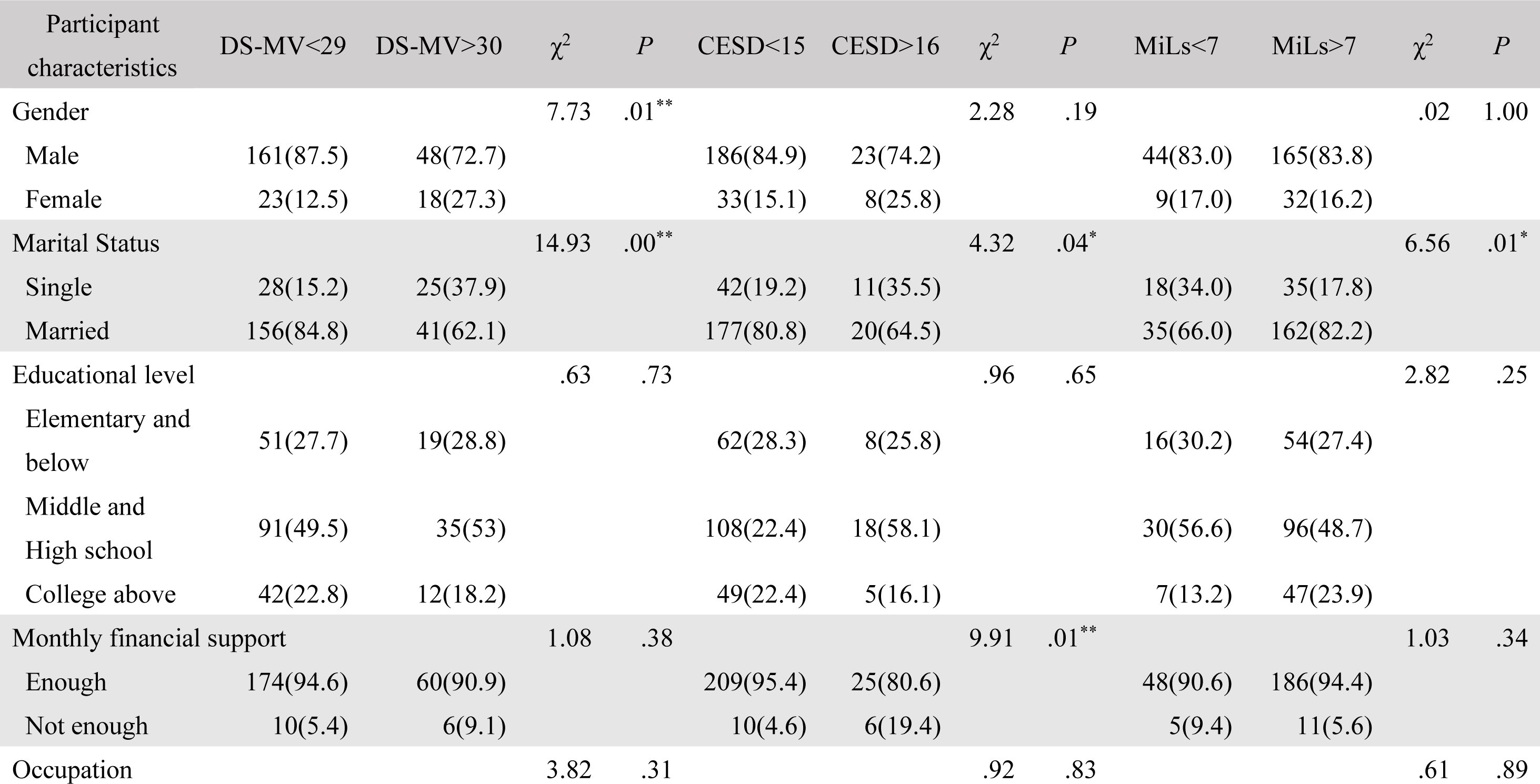
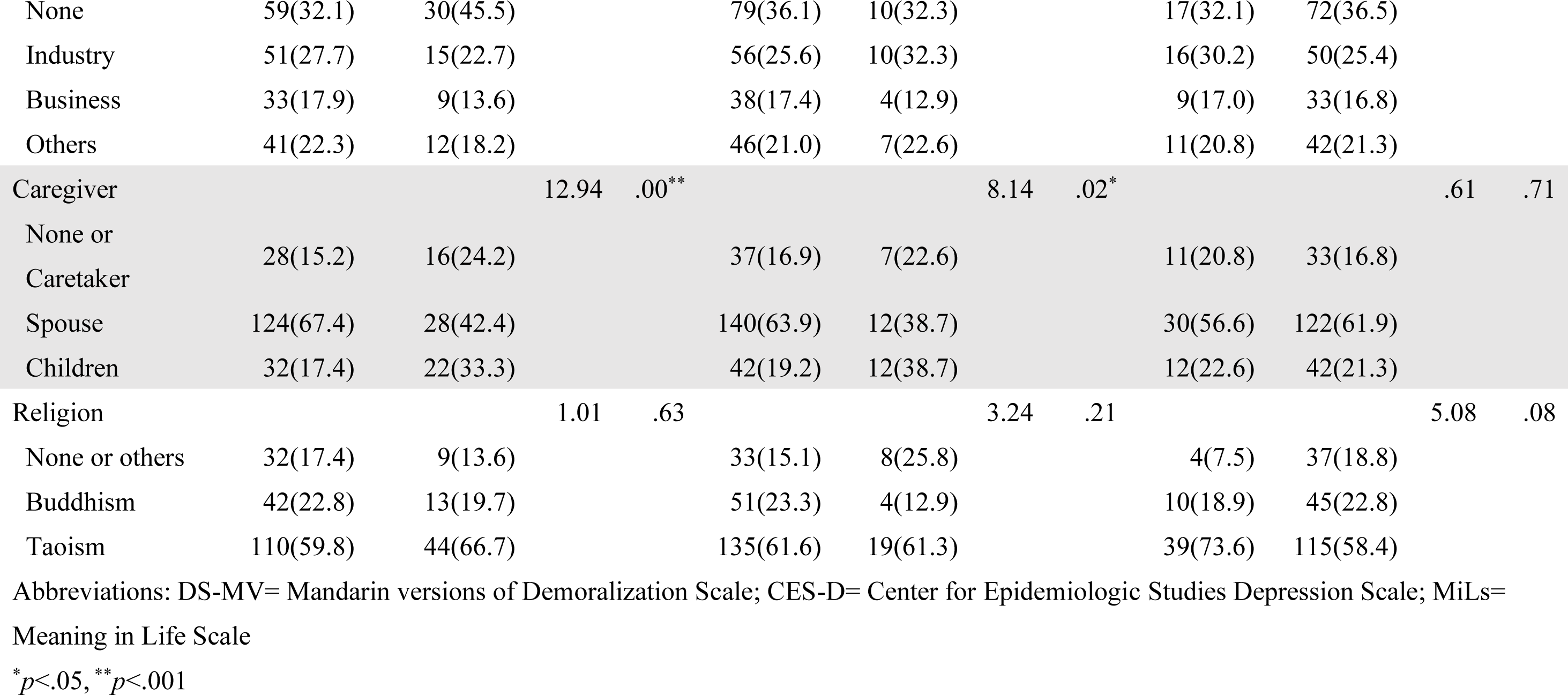
Relationship between demographic factors and demoralization, depression or negative meaning of life in HCC patients (N=250)

Our results showed a significant difference between marital status and DS-MV (*χ^2^*=14.93, *p*<.01), CES-D (*χ^2^*= 4.32, *p*=.04) or MiLs (*χ^2^*=6.56, *p*=.01). The number of single populations with a total score of DS-MV greater than 30 points was 47.2% of single population. Similarly, the number of unmarried patients with a total score of CES-D greater than 16 points was 20.7% of single population. The number of single patients exhibiting negative meaning of life was 33.9% of the single population.

There was a significant difference between gender and DS-MV (*χ^2^*=7.73, *p*<.01); the DS-MV greater than 30 points accounted for 43.9% of the female population. On the other hand, there was no significant difference between gender and CES-D (*χ^2^*=2.28, *p*=.19) or MiLs (*χ^2^*=.02, *p*=1.00).

On the hand, significant difference existed between monthly financial support and CES-D (*χ^2^*=9.91, *p*<.01). The population with CES-D greater than 16 points accounted for 37.5% of patients with financial difficulties.

A significant difference existed between primary caregiver and DS-MV (*χ^2^*=12.94, *p*<.01) or CES-D (*χ^2^*=8.14, *p*=.02). Interestingly, 33.3% of patients with a total score of DS-MV over 30 points or 38.7% of those with a total score of CES-D greater than 16 points were cared by children. However, the main caregivers had no significant relationship with MiLs (*χ^2^*=.61, *p* =.74).

### 4.4 Relationship between clinical factors and demoralization, depression and negative meaning of life in HCC patients

Chi-square test was also used to compare the dichotomous differences in the total scores of cancer stage, cancer recurrence, cancer metastasis, cancer comorbidity (e.g., pain, ascites, fullness) or treatment schemes and demoralization, depression or negative meaning of life in patients with liver cancer **(Table 4)**. The results showed that there was no significant difference between cancer recurrence and DS-MV (*χ^2^*=.40, *p*=.53) or CES-D (*χ^2^*=.67, *p*=.41). There was, however, a significant difference between cancer recurrence and meaning of life score (*χ^2^*=4.52, *p*=.03). The number of participations with a total score of MiLs of less than 7 points was 64.2 % higher than the non-recurrent population.

**TABLE 4.**
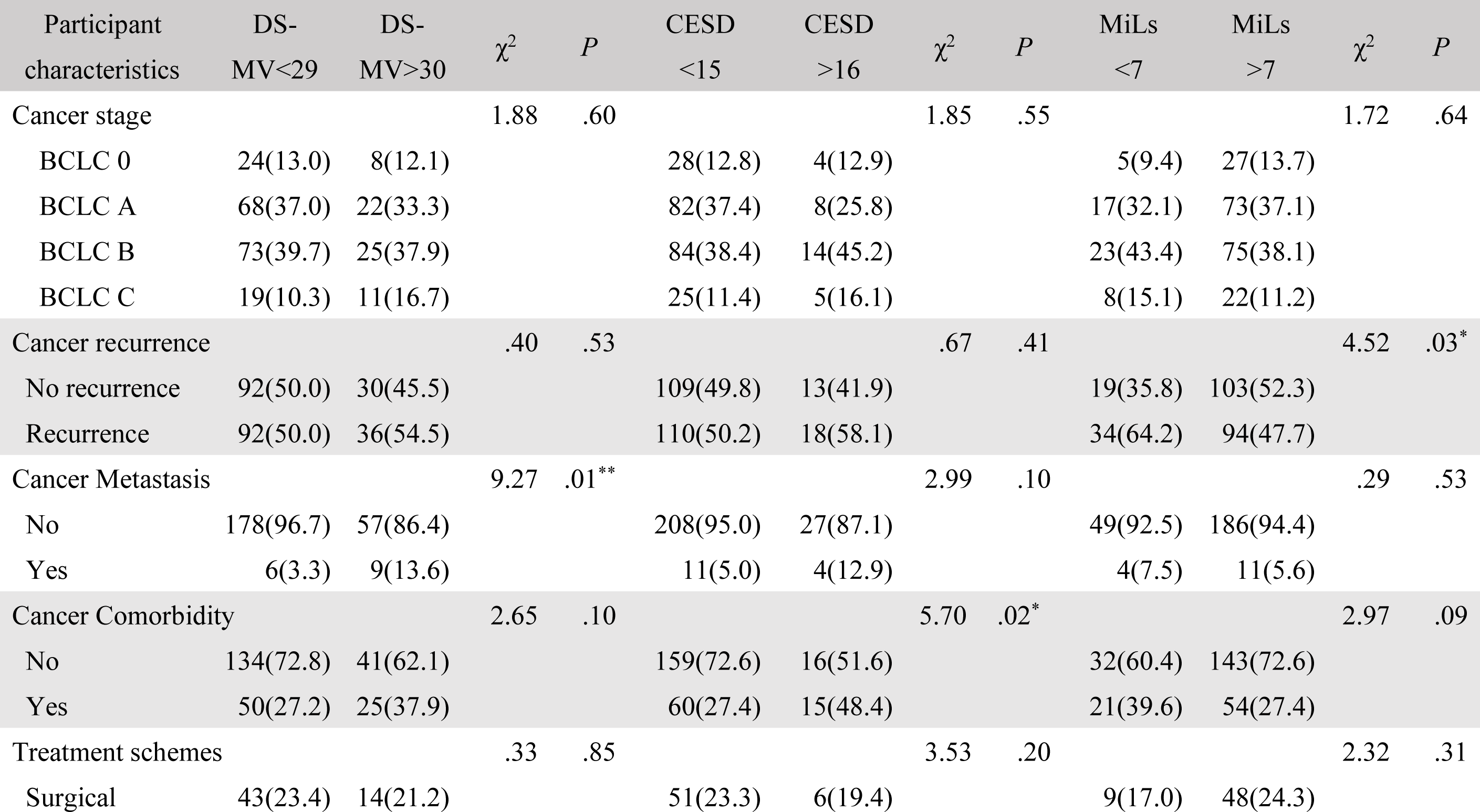
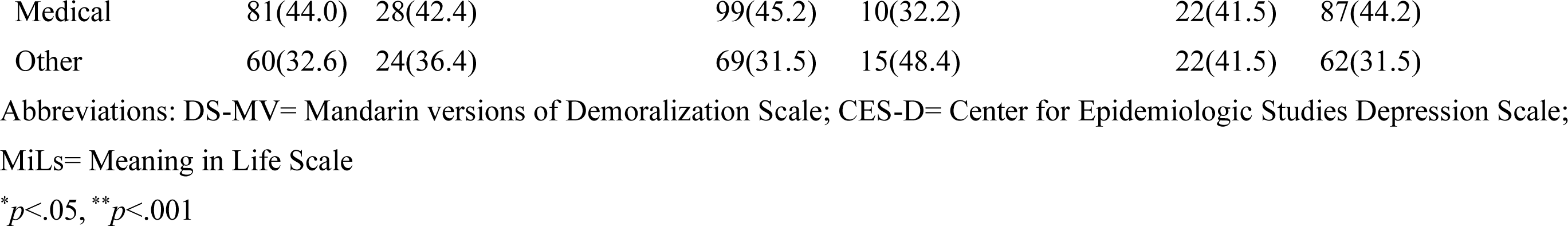
Relationship between clinical factors and demoralization, depression or negative meaning of life in HCC patients. (N=250)

Comorbidity from liver cancer was not significantly related to DS-MV (*χ^2^*=2.65, *p*=.10) and MiLs (*χ^2^*=2.97, *p*=.09), but significantly impacted CES-D (*χ^2^*=5.70, *p*=.02). Participations with cancer comorbidity accounted for 48.8% of the population that exhibited more than 16 points in the CES-D score.

There was a significant difference between cancer metastasis and DS-MV (*χ^2^*=9.27, *p*<.01). Participations with a total score of more than 30 points of DS-MV accounted for 60% of the metastatic population. However, cancer metastasis was not significantly related to CES-D (*χ^2^*=2.99, *p*=.08) and MiLs (*χ^2^*=.29, *p*=.95). Furthermore, there was no significant difference between stage of HCC or treatment schemes and DS-MV, CES-D or MiLs.

### 4.5 Correlation between age or cancer duration and demoralization, depression or negative meaning of life in HCC patients

Finally, Pearson’s correlation was used to analyze the relationship between age or cancer duration and demoralization, depression or negative meaning of life in our HCC patients. Our results showed that age was negatively correlated with the total score of CES-D (*r*=-.13, *p*=.03), and cancer duration was negatively correlated with MiLs (*r*=-.14, *p*=.03). **(Table 5)**.

**TABLE 5.** Correlation between participants age, cancer duration and demoralization, depression or negative meaning of life in HCC patients (N=250)

| Participant characteristics |  | Age | Duration | DS-MV | CES-D | MiLs |
| --- | --- | --- | --- | --- | --- | --- |
| Age | Pearson | 1 |  |  |  |  |
| Duration | Pearson | .18** | 1 |  |  |  |
| DS-MV | Pearson | -.07 | .09 | 1 |  |  |
| CES-D | Pearson | -.13* | =.02 | .53** | 1 |  |
| MiLs | Pearson | -.04 | -.14* | -.52** | -.35** | 1 |

Abbreviations: DS-MV= Mandarin versions of Demoralization Scale; CES-D= Center for Epidemiologic Studies Depression Scale; MiLs= Meaning in Life Scale
\* $p < .05$ , \*\* $p < .001$

## 5. DISCUSSION

The present study was undertaken to assess comprehensively the demographic and clinical factors that may collectively contribute to demoralization, depression and meaning of life in patients with liver cancer in accordance with Asian practice of caregiving. We found that demoralization and depression are positively correlated in HCC patients, and those two mental states are negatively related with meaning of life. We further found, intriguingly, that these three psycho-emotional states in patients with cancer are not uniformly impacted by the eight demographic and six clinical factors that were appraised in this study. We identified that age, gender, marital status, nature of primary caregiver, monthly financial support, cancer metastasis, cancer comorbidity and recurrence or duration are contributing factors to demoralization, depression or meaning of life in HCC patients. On the other hand, our results showed that education, occupation, religion, cancer stage and treatment schemes are unrelated to all three psychological states.

Many studies have shown that patients with cancer who are single are prone to high levels of depression (Arvanitou et al., 2023, Ko et al., 2018; Vehling et al., 2012). Much less work, on the other hand, addressed the relationship between marital status and meaning of life. By showing that patients with cancer who are single were more prone to demoralization, depression and negative meaning of life, this study revealed that marital status is a major contributing factor to all those three psychological states. 78.7% of our study population were married, and 78.7% of those patients exhibited positive meaning of life. This suggests that in Asian culture as seen in Taiwan, there is a tendency for patients with cancer to increase the psychological attachment and dependence on their spouses or partners whose company and support will certainly encourage positive meaning of life. The encouragement and companion of the spouse or partner will enhance the acceptance of patients with cancer treatment, the willingness to maintain good lifestyle and healthy behavior (Gumà & Spijker, 2020; Secinti et al., 2019; Thomas et al., 2017), along with different degrees of spiritual growth, changes in the priority of things, and a clearer understanding of life goals and self-identity (Elekes, 2017). Looking for long-term goals and seeking new experiences are related to the inner sense of existence of an individual (Gumà & Spijker, 2020; Jim et al., 2006). With the secured self-affirmation, it is more likely that patients can overcome the unpleasant experiences associated with malignancy. In short, the more positive the meaning of life, the lower demoralization and depression.

Another intriguing finding in this study is that according to Asian practice of caregiving, a significant relationship existed between the primary caregiver and demoralization or depression. Our results showed that 33.3% of patients with high DS-MV scores and 38.7% of those with high CES-D scores were cared primarily by their children instead of their spouse. The need to rely on the care and assistance of young family members for daily life, the subjective feelings of incompetence and the loss of self-esteem easily lead to negative thoughts of self-perceived burden (Li et al., 2017a). Aggravated by the sense of guilt entrenched in Asian culture, it is likely that the negative emotions such as suicidal tendency and despair that accompany worries of becoming a burden to their younger generation (Bovero et al., 2022; Tang et al. 2020) will evolve to the display of demoralization and depression in patients with liver cancer.

This study showed that both gender and cancer metastasis contribute to demoralization in patients with liver cancer. Whereas the contribution of gender has not been consistent in the literature (Ko et al., 2018; Li et al.,2017a; Vehling et al., 2012), we found that female is one of factors that is associated with demoralization. In the case of cancer metastasis, the concern that malignancy is prolonged and the uncertainty with the length of their lives will together lead patients to irritability and agony that are associated with negative feelings of regret that many things in life remain unaccomplished.

This study also revealed that monthly financial support, cancer comorbidity and age contributed to depression exhibited patients with liver cancer. Studies have shown that cancer symptoms and low socioeconomic will increase the risk of depression in patients with cancer (Cook et al., 2020; Mazor et al., 2019; Su et al., 2019). The quality of life of these patients is often affected by comorbidities such as fatigue, pain and sleep problems, leading to anxiety and depression. These emotional states will be intensified when faced with limited or no source of income, particularly when the patient happens to be the breadwinner of the family and is at a disadvantage during job seeking because of age.

Our results also indicated that both recurrence and duration of cancer were factors that impact on negative meaning of life. One of the common psychological problems (quality of life, mental distress, and worries on financial burden), encountered by patients with cancer is the uncertainty and fear of recurrence despite receiving treatment. Studies have shown that psychological fear is the most obvious when cancer has indeed recurred (Emery et al. 2022, Mardani et al, 2023).

### 5.1 Limitations

Results of this study were obtained from a single investigative population: patients with HCC. Whether our identified demographic and clinical factors that contribute to demoralization, depression and negative meaning of life in patients with other forms of malignancy must therefore be viewed with caution.

### 5.2 Implications for policy and practice

Clinically, the mental states of patients with cancer are not often the primary concern of the nursing staff in performing their daily duties. With our identified demographic and clinical factors that contribute to demoralization, depression and negative meaning of life, clinical nurses should pay more attention to patients with cancer who are single, receive primary care from children, female, aged or face financial difficulties, and alert social workers when signs of psychological disturbance are manifested. Likewise, clinical nurses should be more conscientious of stressful comorbidities that may accompany malignancy, particularly in patients who have a long history of the disease, or when they have been diagnosed of cancer metastasis or recurrence. Psychiatrists should be alerted when signs of demoralization, depression or negative meaning of life are exhibited.

## 6. CONCLUSION

Of the eight demographic and six clinical factors assessed in this comprehensive study on the psychological states of patients with liver cancer, we found that marital status contributes uniformly to demoralization, depression and negative meaning of life. Children as primary caregiver impacts more critically than spouse on both demoralization and depression; gender and cancer metastasis are akin to demoralization; age, monthly financial support and cancer comorbidity contribute to depression; and recurrence and duration of cancer impact on negative meaning of life.

## Data Availability

Send an email to the author to require.

## Acknowledgements

We sincerely thank all patients with liver cancer who participated in this study. We wish particularly to acknowledge Professor Samuel H.H. Chan, Chang Gung Medical Foundation, for his unfailing mentorship on the preparation of this manuscript.

